# Improving on estimates of the potential relative harm to health from using modern ENDS (vaping) compared to tobacco smoking

**DOI:** 10.1101/2020.12.22.20248737

**Authors:** Nick Wilson, Jennifer A Summers, Driss Ait Ouakrim, Janet Hoek, Richard Edwards, Tony Blakely

## Abstract

**Background:** Although the harm to health from electronic nicotine delivery systems (ENDS) compared to smoked tobacco remains highly uncertain, society and governments still need to know the likely range of the relative harm to inform regulatory policies for ENDS and smoking.

**Methods:** We identified biomarkers with specificity of association with different disease groupings e.g., volatile organic compound (VOCs) for chronic obstructive pulmonary disease; and tobacco-specific N’-nitrosamines (TSNAs) and polycyclic aromatic hydrocarbons (PAHs) for all cancers. We conducted a review of recent studies (post January 2017) that compared these biomarkers between people exclusively using ENDS and those exclusively smoking tobacco. The percentage differences in these biomarkers, weighted by study size and adjusted for acrolein from other sources, were used as a proxy for the assumed percentage difference in disease harm between ENDS and smoking. These relative differences were applied to previously modelled estimates of smoking-related health loss (in health-adjusted life-years; HALYs).

**Results:** The respective relative biomarker levels (ENDS vs smoking) were: 28% for respiratory diseases (five results, three studies); 42% for cancers (five results, four studies); and 35% for cardiovascular (seven results, four studies). When integrated with the HALY impacts by disease, the overall harm to health from ENDS was estimated to be 33% that of smoking.

**Conclusions:** This analysis, suggests that the use of modern ENDS devices (vaping) could be a third as harmful to health as smoking in a high-income country setting. But this estimate is based on a limited number of biomarker studies and is best be considered a likely upper level of ENDS risk given potential biases in our method (i.e., the biomarkers used being correlated with more unaccounted for toxicants in smoking compared to with using ENDS).

## INTRODUCTION

The extent to which public health agencies and governments should restrict or support access to electronic nicotine delivery systems (ENDS), either as a means of quitting smoking or for harm reduction (relative to smoking) if long-term nicotine use persists, is controversial internationally. Previous estimates such as around 5% of the relative harm to health of ENDS use vs tobacco smoking [1, 2], are not disease-specific and have been critiqued, partly because they rely mainly on comparisons of emission levels from ENDS devices and tobacco smoking [3, 4], rather than studies of biomarkers or health outcomes. Studies of the relative harm of aerosol vs smoke are very limited because the relationships between emissions and biological outcomes remain unclear, and because ENDS users and smokers have different inhalation patterns. Other recent provisional review work on ENDS by a UK Government group makes only vague comments about ENDS having a “substantially lower” risk of adverse health effects compared to smoked cigarettes [5]. A recent review of six studies reported that former smokers who transitioned to e-cigarettes “showed ∼ 40% lower odds of respiratory outcomes compared to current exclusive smokers” [6]. However, the authors also noted that “switching from smoking to e-cigarette[s] does not appear to significantly lower odds of cardiovascular outcomes.” Overall, many limitations affect interpretation of these studies, as five were cross-sectional and only one was longitudinal.

Changes in ENDS technology also suggest the likely relative harm may change over time as device design and quality control of manufacturing processes of the e-liquid and nicotine salt solutions evolve. Dynamic product development and manufacturing suggest that estimates of relative harm should be based on data from recently conducted studies of ENDS.

Despite uncertainty about the health effects of ENDS use, societies and policy-makers still need to make policy on how they are regulated. Their decision-making often relies on modelling studies (e.g., as per these ones: [7-18]), which require up-to-date and credible estimates of harm arising from ENDS use relative to tobacco smoking. The most recent of these modelling studies we identified, used a relative harm range from 5% to 20% [18], but did not provide a detailed justification for these values. Improved quantification of the relative harm should improve policy-making and assist smokers deciding whether it is better to switch to ENDS use or continue trying to quit all nicotine products.

In the absence of adequate long-term epidemiological data on the health effects of ENDS use, studies comparing levels of biomarkers associated with the occurrence of adverse health outcomes between exclusive smokers and ENDS users may provide more valid comparisons of relative health impacts than reviews using mainly emissions-based data. That is, biomarkers are likely to more closely represent the actual exposure of organs and tissues than will emissions-based studies. Select biomarkers for smoking-related toxicants are associated with key adverse health outcomes in smokers (Table 1), even though there is “variation in exposure due to differences in smoke composition across brands and to inherent variability among smokers” [19]. Therefore in this study we conducted a review of relevant and recent biomarker data with the aim of producing an updated estimate for relative harm of ENDS use compared to smoking.

**Table 1:** Relationship between health impacts from smoking and key biomarkers for toxicants

| Health loss from smoking | Biomarkers | Sources, comment |
| --- | --- | --- |
| Chronic obstructive pulmonary disease (COPD) | Volatile organic compounds (VOCs) e.g., acrolein, crotonaldehyde | The WHO [20], considers these agents to be hazardous with acrolein considered to be: "an intense irritant, is toxic to lung cilia and has been proposed as a lung carcinogen". Similarly, "crotonaldehyde is a potent irritant and a weak hepatocarcinogen and forms DNA adducts in the human lung." |
| All cancers | Tobacco-specific N'-nitrosamines (TSNAs) | The WHO [20], notes that two TSNAs, "NNK and NNN, are probably responsible for cancers of the lung, pancreas, oral cavity and oesophagus in tobacco users". "Both have been classified as human carcinogens by working groups at [International Agency for Research on Cancer] IARC." (See Table 3 regarding NNK and NNN and the full terms). |
|  | Polycyclic aromatic hydrocarbons (PAHs) | The WHO [20], notes that: "many PAHs are potent carcinogens or toxicants in laboratory animals (57), and many are present in cigarette smoke, including the prototypic PAH benzo[a]pyrene, classified as a human carcinogen" by a working group convened by the IARC. |
| Cardiovascular disease | Carbon monoxide (CO) | The WHO [20], states that: "CO is a well established cardiovascular toxicant, which competes with oxygen for binding to haemoglobin. In smokers, it is considered to reduce oxygen delivery, cause endothelial dysfunction and promote the progression of atherosclerosis and other cardiovascular diseases". A US Surgeon General's Report also states that: "the mechanisms by which CO may contribute to acute cardiovascular events are well characterized" [21].<br>The WHO [20] reports that: "cardiovascular tissues appear to be |
|  | Acrolein | particularly sensitive to the toxic effects of acrolein". A review on this association has also been published [22]. |

## METHODS

To summarise, our method used the following three steps:

1. We identified biomarkers with specificity of association with different disease groupings: volatile organic compound (VOCs) for chronic obstructive pulmonary disease (COPD); tobacco-specific N’-nitrosamines (TSNAs) and polycyclic aromatic hydrocarbons (PAHs) for all cancers; and carbon monoxide and the VOC, acrolein, for cardiovascular disease (CVD).
2. We conducted a review of recent studies (published and with data collected after January 2017) that compared these biomarkers (in blood, urine and exhaled breath) between people exclusively using ENDS and those exclusively smoking tobacco, determining the percentage difference in these biomarkers between ENDS users and smokers.
3. These percentage differences in biomarkers were assumed to reflect the percentage difference in disease harm, and were applied to previously modelled estimates of smoking-related health loss (in health-adjusted life-years; HALYs) to produce disease-group-specific and overall estimates of health loss for ENDS use versus tobacco smoking.

### Linking key biomarkers with categories of health loss

For this process we relied on a recent key World Health Organization Report detailing recent biomarker research [20], supplemented with other key literature. The details are in Table 1.

### Literature searches to identify relevant biomarker studies of the differences in biomarkers between ENDS users and smokers

Searches of the peer-reviewed literature were conducted using PubMed and Google Scholar on 1 September 2020 using the names of relevant toxicants and biomarkers as listed in a FDA Review document [23] (see Supplementary Information for further details and a PRISMA flow diagram: Supplementary Tables 1 to 3).

We considered studies published from 1 January 2017 to 1 September 2020. To be included, studies had to compare exclusive ENDS users to exclusive tobacco smokers for the relevant biomarker in either urine, blood or exhaled air. Studies based on data collected prior to 1 January 2017 were excluded as we aimed to focus on the most recent ENDS devices and to increase the probability that relatively modern quality control measures were being used by the manufacturers of devices and e-liquids. Additional information describing the methodology of the review are provided in the Supplementary Information.

### Health loss by disease categories

We used the results of a tobacco epidemiology and control modelling study [24], which has been extensively used for modelling tobacco control interventions [25-30]. This model allows for the examination of tobacco control interventions on health across the whole life course in the population. It identifies HALYs gained from tobacco control interventions for preventing the following four disease groupings: COPD, cancers (a grouping of 12 tobacco-associated cancers), CVD, and lower respiratory tract infection [24]. The results for these different condition groups are detailed in Table 2.

**Table 2:** Health impacts by disease group as a result of modelling a tobacco control intervention (tobacco tax increases) at a national level [24]

| Health condition / condition group | Proportion of HALYs gained* from preventing uptake and promoting quitting of smoking (undiscounted) |
| --- | --- |
| Chronic obstructive pulmonary disease (COPD) | 48.9% |
| Cancers (12 types**) | 28.3% |
| CVD (coronary heart disease and stroke) | 22.4% |
| Lower respiratory tract infection | 0.4% |
| **Total** | **100%** |

\* Specifically from a tobacco tax intervention in New Zealand (a 10% per annum increase in tobacco tax from 2011 to 2031 that impacts on both increasing quitting and reducing youth uptake), and values from Table S6 in the S2 Supplementary Text of Blakely et al [24]. The HALYs are for the 2011 population over the remainder of their lifespans. Therefore, many of the health gains are decades into the future. American Cancer Society's Cancer Prevention Study II (CPS II) relative risks are used in this particular analysis (with results being similar to relative risks calculated from New Zealand studies).
\*\* Cancers in descending order of importance as per the BODE<sup>3</sup> preferred model: Lung cancer (26.0%), bladder cancer (1.3%), mouth and oropharyngeal cancer (0.9%), oesophageal cancer (0.9%), liver cancer (0.9%), pancreatic cancer (0.6%), stomach cancer (0.6%), kidney cancer (0.2%), cervical cancer (0.2%), thyroid cancer (0.0%), endometrial cancer (-0.2%), melanoma (-0.8%).

### Integration of the relative biomarker results with the health impact results

Our final analysis integrated the results of the relative biomarker levels (taking the weighted mean result with weighting based on study participant total numbers), and the relative HALY impacts from the epidemiological model (Table 2). But as one particular toxicant, acrolein, has other major sources (e.g., air pollution) we further adjusted the relative harm values for acrolein using data from the largest relevant study we found in the literature (Alwis et al [31], see Supplementary Table 5). For this adjustment we assumed that other sources of acrolein (diet, air pollution etc) were non-differential between ENDS users and smokers.

For smokers who become ENDS users, the washout period used by the study may be important. That is for the biomarker NNAL, the half-life in the human body is 10 to 18 days [32]. Our analysis did include two cross-sectional studies and one experimental study with eight weeks of follow-up using NNAL. But given this half-life issue, we excluded results for NNAL from one short-term experimental study lasting five days [33], where there would have been inadequate time for NNAL levels to have fully equilibrated with the transition to exclusive use of ENDS. But for all other biomarkers considered, half-lives were under 10 hours and so this need for a long washout period was not relevant (see Table 3 footnotes).

**Table 3:**
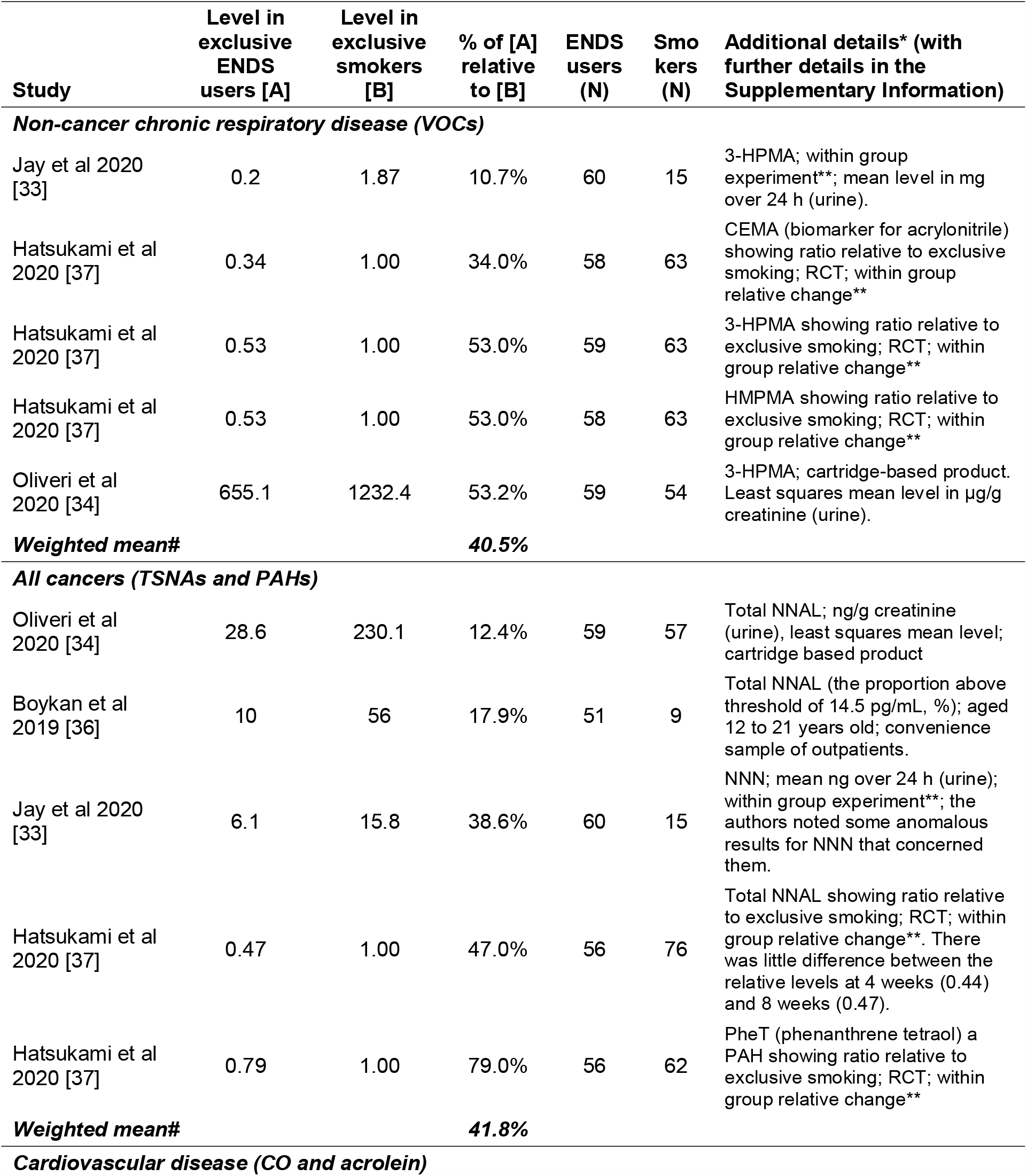
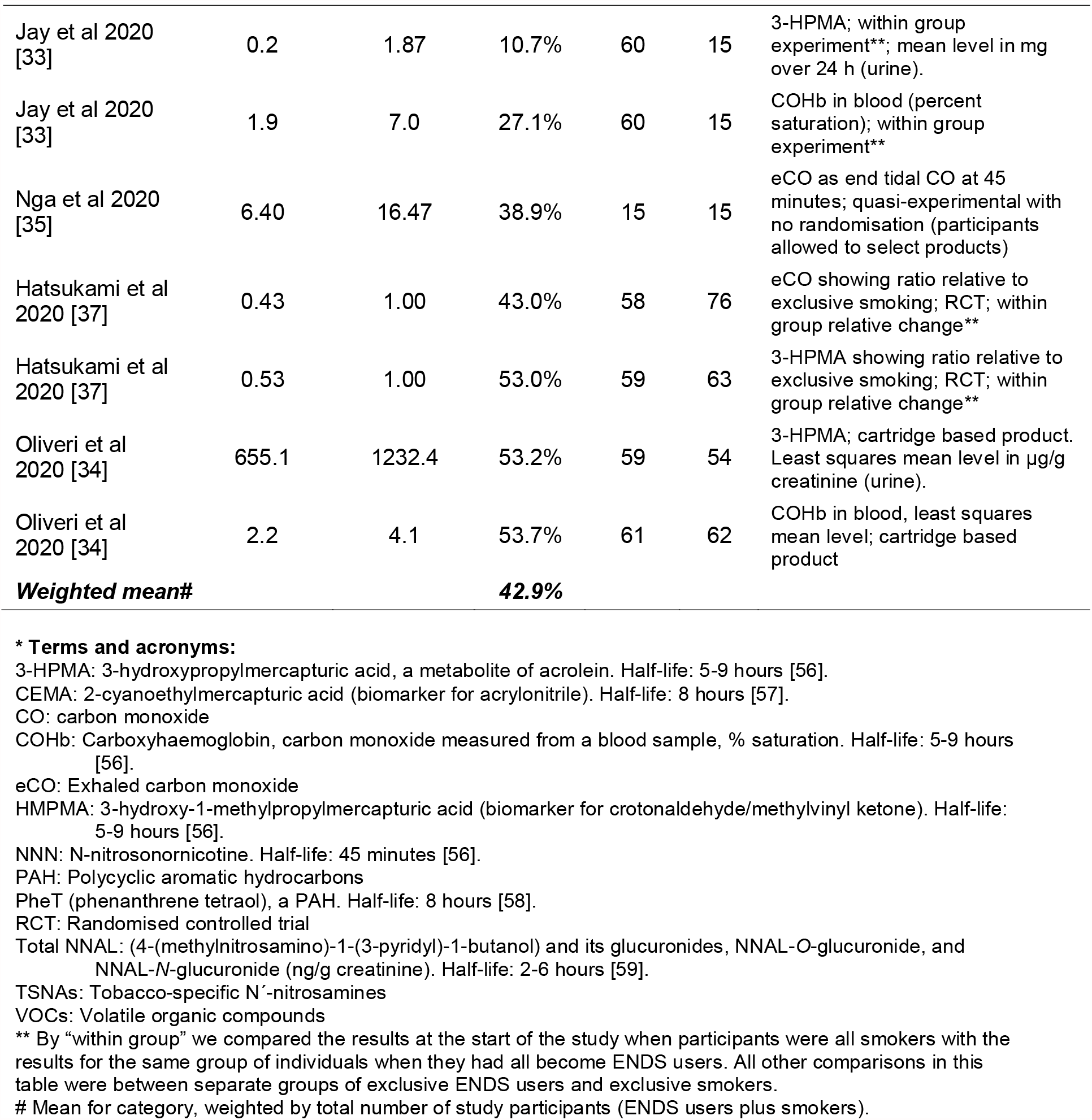
Results from the recent biomarker studies identified involving use of modern ENDS products (and data on exclusive ENDS use and exclusive smoking with data collection since 1 January 2017)

| Study | Level in<br>exclusive<br>ENDS<br>users [A] | Level in<br>exclusive<br>smokers<br>[B] | % of [A]<br>relative<br>to [B] | ENDS<br>users<br>(N) | Smokers<br>(N) | Additional details* (with<br>further details in the<br>Supplementary Information) |
| --- | --- | --- | --- | --- | --- | --- |
| **Non-cancer chronic respiratory disease (VOCs)** |  |  |  |  |  |  |
| Jay et al 2020<br>[33] | 0.2 | 1.87 | 10.7% | 60 | 15 | 3-HPMA; within group<br>experiment**; mean level in mg<br>over 24 h (urine). |
| Hatsukami et al<br>2020 [37] | 0.34 | 1.00 | 34.0% | 58 | 63 | CEMA (biomarker for acrylonitrile)<br>showing ratio relative to exclusive<br>smoking; RCT; within group<br>relative change** |
| Hatsukami et al<br>2020 [37] | 0.53 | 1.00 | 53.0% | 59 | 63 | 3-HPMA showing ratio relative to<br>exclusive smoking; RCT; within<br>group relative change** |
| Hatsukami et al<br>2020 [37] | 0.53 | 1.00 | 53.0% | 58 | 63 | HMPMA showing ratio relative to<br>exclusive smoking; RCT; within<br>group relative change** |
| Oliveri et al<br>2020 [34] | 655.1 | 1232.4 | 53.2% | 59 | 54 | 3-HPMA; cartridge-based product.<br>Least squares mean level in µg/g<br>creatinine (urine). |
| **Weighted mean#** |  |  | **40.5%** |  |  |  |
| **All cancers (TSNAs and PAHs)** |  |  |  |  |  |  |
| Oliveri et al<br>2020 [34] | 28.6 | 230.1 | 12.4% | 59 | 57 | Total NNAL; ng/g creatinine<br>(urine), least squares mean level;<br>cartridge based product |
| Boykan et al<br>2019 [36] | 10 | 56 | 17.9% | 51 | 9 | Total NNAL (the proportion above<br>threshold of 14.5 pg/mL, %); aged<br>12 to 21 years old; convenience<br>sample of outpatients. |
| Jay et al 2020<br>[33] | 6.1 | 15.8 | 38.6% | 60 | 15 | NNN; mean ng over 24 h (urine);<br>within group experiment**; the<br>authors noted some anomalous<br>results for NNN that concerned<br>them. |
| Hatsukami et al<br>2020 [37] | 0.47 | 1.00 | 47.0% | 56 | 76 | Total NNAL showing ratio relative<br>to exclusive smoking; RCT; within<br>group relative change**. There<br>was little difference between the<br>relative levels at 4 weeks (0.44)<br>and 8 weeks (0.47). |
| Hatsukami et al<br>2020 [37] | 0.79 | 1.00 | 79.0% | 56 | 62 | PheT (phenanthrene tetraol) a<br>PAH showing ratio relative to<br>exclusive smoking; RCT; within<br>group relative change** |
| **Weighted mean#** |  |  | **41.8%** |  |  |  |
| **Cardiovascular disease (CO and acrolein)** |  |  |  |  |  |  |
| Jay et al 2020<br>[33] | 0.2 | 1.87 | 10.7% | 60 | 15 | 3-HPMA; within group<br>experiment**; mean level in mg<br>over 24 h (urine). |
| Jay et al 2020<br>[33] | 1.9 | 7.0 | 27.1% | 60 | 15 | COHb in blood (percent<br>saturation); within group<br>experiment** |
| Nga et al 2020<br>[35] | 6.40 | 16.47 | 38.9% | 15 | 15 | eCO as end tidal CO at 45<br>minutes; quasi-experimental with<br>no randomisation (participants<br>allowed to select products) |
| Hatsukami et al<br>2020 [37] | 0.43 | 1.00 | 43.0% | 58 | 76 | eCO showing ratio relative to<br>exclusive smoking; RCT; within<br>group relative change** |
| Hatsukami et al<br>2020 [37] | 0.53 | 1.00 | 53.0% | 59 | 63 | 3-HPMA showing ratio relative to<br>exclusive smoking; RCT; within<br>group relative change** |
| Oliveri et al<br>2020 [34] | 655.1 | 1232.4 | 53.2% | 59 | 54 | 3-HPMA; cartridge based product.<br>Least squares mean level in µg/g<br>creatinine (urine). |
| Oliveri et al<br>2020 [34] | 2.2 | 4.1 | 53.7% | 61 | 62 | COHb in blood, least squares<br>mean level; cartridge based<br>product |
| **Weighted mean#** |  |  | **42.9%** |  |  |  |

**\* Terms and acronyms:**
3-HPMA: 3-hydroxypropylmercapturic acid, a metabolite of acrolein. Half-life: 5-9 hours [56].
CEMA: 2-cyanoethylmercapturic acid (biomarker for acrylonitrile). Half-life: 8 hours [57].
CO: carbon monoxide
COHb: Carboxyhaemoglobin, carbon monoxide measured from a blood sample, % saturation. Half-life: 5-9 hours [56].
eCO: Exhaled carbon monoxide
HMPMA: 3-hydroxy-1-methylpropylmercapturic acid (biomarker for crotonaldehyde/methylvinyl ketone). Half-life: 5-9 hours [56].
NNN: N-nitrosornicotine. Half-life: 45 minutes [56].
PAH: Polycyclic aromatic hydrocarbons

## RESULTS

### Identified biomarker studies

The results of our literature search and study selection process are shown in a PRISMA flow diagram (see Supplementary Information). Out of the 584 identified studies, five met our inclusion criteria by having appropriate comparison groups and data on contemporary ENDS devices (since 1 January 2017): Oliveri et al 2020 [34], Jay et al 2020 [33], Nga et al 2020 [35], Boykan et al 2019 [36], and Hatsukami et al 2020 [37].

Two were experimental studies, one quasi-experimental and two were cross-sectional. In terms of study quality, two of the five studies were funded by commercial interests in ENDS use/tobacco [33, 34]. Nevertheless, a particular advantage of one of these was that it kept users in a controlled environment (albeit only for five days) [33], which may have reduced the risks of contamination via undeclared dual use of ENDS use and tobacco products, as well as exposure to secondhand smoke/ENDS aerosol from others. Other study design limitations of the included studies included involving narrow demographic groups (e.g., those aged 12 to 21 years [36]) and allowing participants to select products themselves [35], as opposed to being randomised. In the two cross-sectional studies, the authors had to rely on self-reporting as to participants being “exclusive” smokers or “exclusive” ENDS users. Further details on all these studies, including the specific ENDS products used are presented in Supplementary Table 4.

Many of the identified biomarker studies excluded from the analysis had collected data before our cut-off period of prior to January 2017 (e.g., [38-48]). Furthermore, some more recent studies did not involve appropriate comparison groups i.e., did not compare exclusive ENDS users with exclusive tobacco smokers (e.g., [49-53]). We excluded one study [54] because participants had occupational exposure to volatile chemicals (i.e., workers in a chemical factory). Another study on exhaled VOCs [55], was excluded because it was unclear whether the exhaled chemicals could be considered to be solely biomarkers or whether these also involved un-metabolised aerosol/smoke from recent inhalation of the products.

### Biomarker results by disease categories

Table 3 shows the results of the relative levels of the selected biomarkers. The mean results weighted by study size and adjusted for acrolein from other sources were: 28% for respiratory diseases (five results, three studies); 42% for cancers (five results, four studies); and 35% for CVD (seven results, four studies).

### Integrated analysis of biomarkers and health loss

When integrated with the HALY impacts from a modelled tobacco control intervention, by disease grouping, and with downward adjustments due to acrolein from other sources, the overall harm to health from ENDS was estimated at 33.2% that of smoking (Table 4).

**Table 4:**
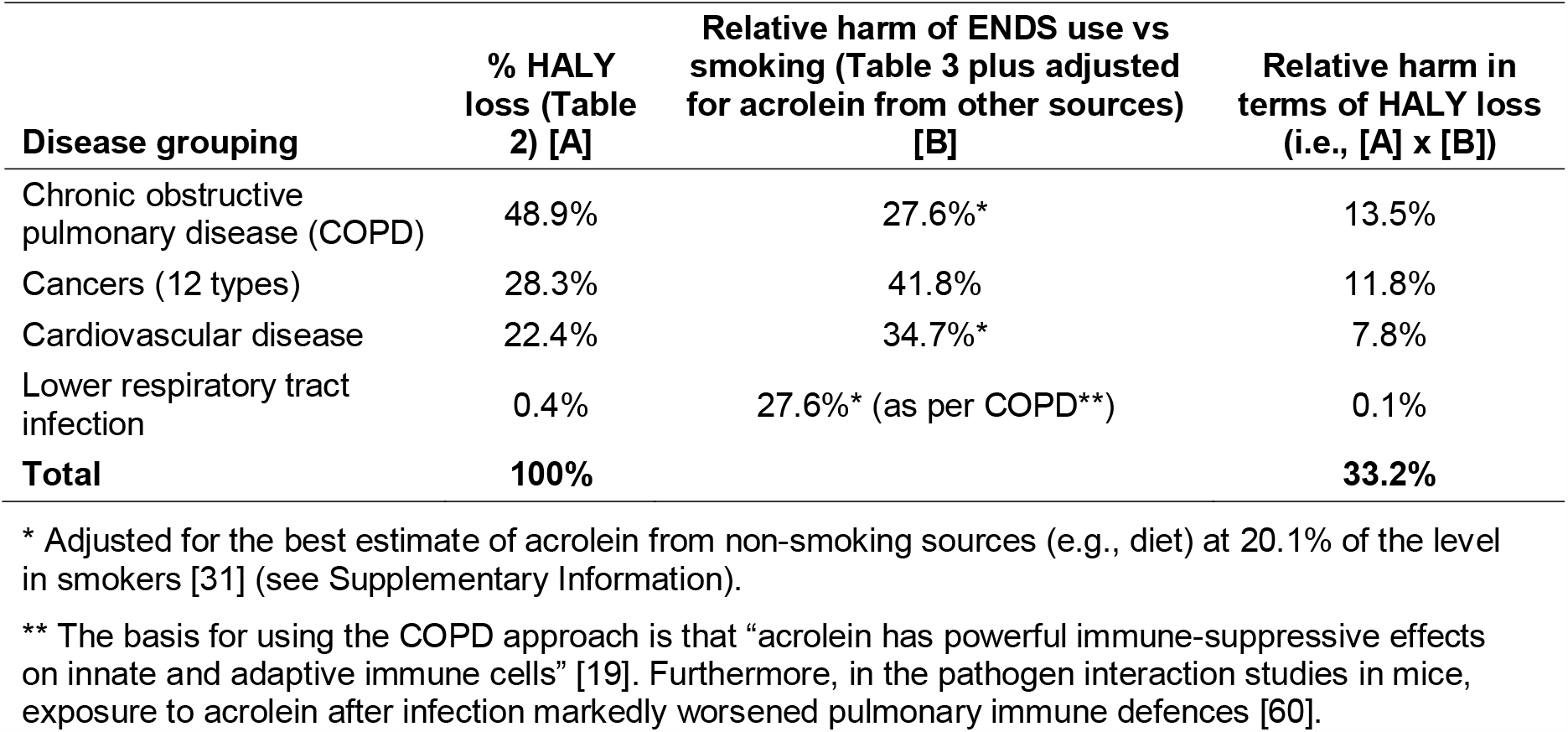
Integrated analysis of the relative harm from using modern ENDS devices relative to smoking tobacco in terms of health loss in HALYs by disease grouping

## DISCUSSION

This analysis combined recent biomarker data from ENDS use (relative to smoking) with modelled smoking health loss data to produce an overall estimate of relative harm for ENDS use for four of the main disease groupings caused by smoking tobacco. Our method estimated that the harm associated with modern ENDS was 33% of the harm associated with tobacco smoking. This value is higher than previously suggested (e.g., at around 5% [1, 2]) and the range of relative harm values (5% to 20%) [18], used in the most recent modelling study we identified.

This 33% estimate should be considered a likely upper level given potential biases in our method. A key such potential bias is that there may be more unmeasured toxicants correlated with the biomarkers we measured in smokers compared to ENDS users. Our reasoning is as follows. There are over 7000 chemicals in tobacco smoke, hundreds of which are toxic and around 70 which cause cancer [61]. In contrast, the best estimate we identified to date was of “over 80” chemicals in ENDS aerosol [62]. Now assume the “representative” toxicant biomarker we used in our analysis for each disease did not capture all the causal mechanisms of tobacco smoking with disease (either directly or by correlation), and that the occurrence of other correlated toxicants and mechanisms from ENDS use is less than with smoking tobacco (as suggested by the numbers of chemicals above), our method will likely have overestimated the percentage of harm from ENDS use as compared to smoking. For example, let us assume that acrolein is causally responsible for 60% of excess COPD due to smoking but 80% of excess COPD due to ENDS use (while ignoring toxicants and mechanisms correlated to acrolein). Then our method assumes all of the COPD variation can be explained by acrolein (100%/60% = 1.67 times overestimated). If we then apply it to ENDS use, this would lead to a net 1.33-fold over-estimate (80% times 1.67). The net bias will vary further due to toxicants correlated to acrolein in ENDS aerosol. Indeed, in the unlikely circumstance of there being many other correlated toxicants with ENDS use (or a few very potent correlated toxicants), our method may actually under-estimate the harm from ENDS – but we believe this to be very unlikely.

To further illustrate the potential relevance of toxicants we have not included in our analysis, there is a systematic review [63], which reported that: “Most metal/metalloid levels found in biosamples of e-cigarette users were similar or higher than levels found in biosamples of conventional cigarette users, and even higher than those found in biosamples of cigar users.” We also did not identify any biomarker data relating to formaldehyde, which is commonly detected in ENDS products [64]. Similarly, we did not include fine particulates from ENDS, which may play a role in CVD [65]. Flavourings also differ between ENDS products, and some may have unique lung damaging effects [66]. We also did not include studies of biomarkers of tissue/physiological impact (e.g., respiratory lung inflammation, platelet aggregation etc) given the lack of validation relative to chronic disease outcomes and/or because of the lack of direct comparisons between smoking and ENDS use.

Other limitations of our work include the following:

- Another reason why we may have over-estimated the relative harm of ENDS is that some “exclusive” ENDS users may have been “ex-smokers”, some of whom may have still been smoking. This would result in an underestimate of the true difference in exposure between the groups. This may have been less likely in the experimental studies as each included a measure expected to reduce the likelihood of this bias operating. These measures were use of incentives for compliance [37], confinement to maximise restriction to the allocated product type [33], and screening for evidence of continued smoking [35, 36]. Nevertheless, although there was variation in the findings between studies, the two cross-sectional studies [34, 36], did not report a systematically higher level of biomarkers than the experimental studies, as might be expected if contamination by unreported continued smoking among exclusive ENDS users were greater in these studies.
- The disease categories we analysed only covered four main groupings of tobacco-related disease, but omitted less major ones. For example, there is evidence that smoking causes diabetes and increases the risk of tuberculosis, various eye diseases and immune system disorders such as rheumatoid arthritis [19]. Furthermore, some toxicants in ENDS products (e.g., acrolein) have also been associated with increasing the risk of diabetes [20].
- Within the disease categories we did not differentially weight particular toxicants by their likely importance in disease causation e.g., TSNAs vs PAHs in the “all cancers” grouping. While some work on relative prioritisation has been done (e.g., in tobacco-industry funded research [67]), this work does not appear to be comprehensive enough to produce reliable rankings. Furthermore, we did not consider non-linear dose response relationships. For example, lower levels of smoking intensity and second-hand smoke exposure have disproportionately higher relative risks for CVD than would be expected if the dose-response relationship was linear [19]. These non-linear relationships could mean that we have partly under-estimated the relative harm from toxicants that ENDS users are exposed to and that are associated with cardiovascular disease.
- The biomarker studies represent points in time in the long-term trajectory of ENDS use by individuals and within populations, and include diverse brands and product types (of both ENDS products and comparative tobacco brands). Trajectories of ENDS use and smoking may diverge further in the future. For example, smoked tobacco products have changed little over many decades and we suspect that many smokers will continue smoking long term at approximately the same intensity. However, we are less certain for ENDS use. ENDS users may be more or less likely to continue ENDS use long term compared to smokers. There may also be future changes to ENDS technology and usage patterns that affect exposure levels among ENDS users (e.g., based on changes in relative nicotine levels, or potential delineation of smokefree and vapefree areas, or if public tolerance of ENDS increases relative to smoking, or if ENDS products evolve further).
- More specifically, two of the biomarker studies involved short-term use of ENDS (i.e., for only five days [33], or just a matter of hours [35]). Usage patterns among short-term users may have differed from those exhibited by more experienced ENDS users and this could have impacted on their biomarker measurements.
- Some of the included biomarker studies had limitations and potential biases in their assessment of specific biomarkers among ENDS users. For example, while our analysis adjusted for other sources of acrolein (e.g. dietary sources), we did not have the data to adjust other biomarkers by exposure to secondhand smoke (or secondhand exposure to aerosol from ENDS). Nevertheless, such exposures are likely to be relatively minor given evidence that NNAL levels in non-smokers are typically 1-5% those of smokers (due to exposure to second-hand smoke) [23]. Also, although one study included results for a PAH [37], which has other sources (e.g., cooking emissions, vehicle emissions, and industrial air pollution [68]), this study had the advantages of being a randomised trial, thus such exposures should have been non-differential. But this study was still suboptimal for our purposes in terms of not also measuring PAH in a control group (non-ENDS using and non-smoking), but it did show that PAH levels declined significantly in those switching to exclusive vaping.
- Two [33, 34] of the five biomarker studies used in our main analysis were industry-funded. Given evidence that this conflict of interest is strongly associated with results favourable to the tobacco industry, indicating no harm of ENDS, further caution is required [69].

### Potential research implications

The high level of uncertainty of the relative harm of ENDS use compared to smoking highlights the need to develop a much stronger evidence base. Agencies that fund research should therefore commission further studies that measure a wider range of biomarkers in long-term exclusive ENDS users and long-term exclusive smokers, in addition to long-term epidemiological studies that measure health outcomes. There is also a need for studies on the full range of ENDS products (and wide range of tobacco brands) and for regularly repeated studies given the rapid rate of technological development with ENDS to identify if new devices/e-liquids change biomarker levels. Also, given the limitations around the range of biomarkers in our analysis, additional biomarkers studied should include: PAHs, aromatic amines, acyclic amines, fine particulates, heavy metals and dysregulated metabolites [70]. There may also be a need for expert elicitation exercises involving toxicologists and epidemiologists to estimate the uncertainty ranges. In the interim, however, modelling work done to inform the regulation of ENDS and smoking, should probably use wide uncertainty intervals (as we have ourselves done [71]).

## Conclusions

This analysis suggests that the use of modern ENDS devices (vaping) could be up to a third as harmful to health as smoking in a high-income country setting. This is best considered a likely upper level given the potential biases in our method (i.e., the biomarkers used being correlated with more unaccounted for toxicants in smoking compared to with using ENDS).

## Data Availability

All the relevant data is in the manuscript and Supplementary Information file.

## List of abbreviations

3-HPMA: 3-hydroxypropylmercapturic acid
CEMA: 2-cyanoethylmercapturic acid
CO: carbon monoxide
COHb: Carboxyhaemoglobin
COPD: Chronic obstructive pulmonary disease
CVD: Cardiovascular disease
eCO: Exhaled carbon monoxide
ENDS: Electronic nicotine delivery systems
HALY: Health-adjusted life-years
HMPMA: 3-hydroxy-1-methylpropylmercapturic acid
NNAL: (4-(methylnitrosamino)-1-(3-pyridyl)-1-butanol)
NNN: N-nitrosonornicotine
PAH: Polycyclic aromatic hydrocarbons
PheT: Phenanthrene tetraol
RCT: Randomised controlled trial
TSNAs: Tobacco-specific N’-nitrosamines
VOCs: Volatile organic compounds
WHO: World Health Organization

## Acknowledgements

We thanks Sophie Braznell from the University of Bath, United Kingdom, for helpful comments on the manuscript.

## Author contributions

This work was designed by NW and TB. JH obtained the funding. Literature searches were conducted by JS, DAQ, and NW. Data analysis and writing the first draft was by NW. All authors assisted with revising manuscript drafts.

## Availability of data and materials

All data are contained in the main manuscript and in the Supplementary Information file.

## Ethics approval and consent to participate

Not applicable.

## Consent for publication

Not applicable.

## Supplementary Information

### Literature Search Criteria, Database Search Terms and PRSIMA diagram

We used all the identified biomarkers for tobacco smoking in a FDA Review document [1] (except for those relating to nicotine and tobacco alkaloids), for inclusion in our literature search. The search criteria and specific terms used are shown in Supplementary Tables 1 to 3 below and Supplementary Figure 1 is of the PRISMA diagram of the process. Two reviewers screened one database each. After initial selection based on titles and abstracts, both reviewers compared selected articles and screened full-text articles against the selection criteria. The third reviewer screened the final selection of articles.

**Supplementary Table 1:**
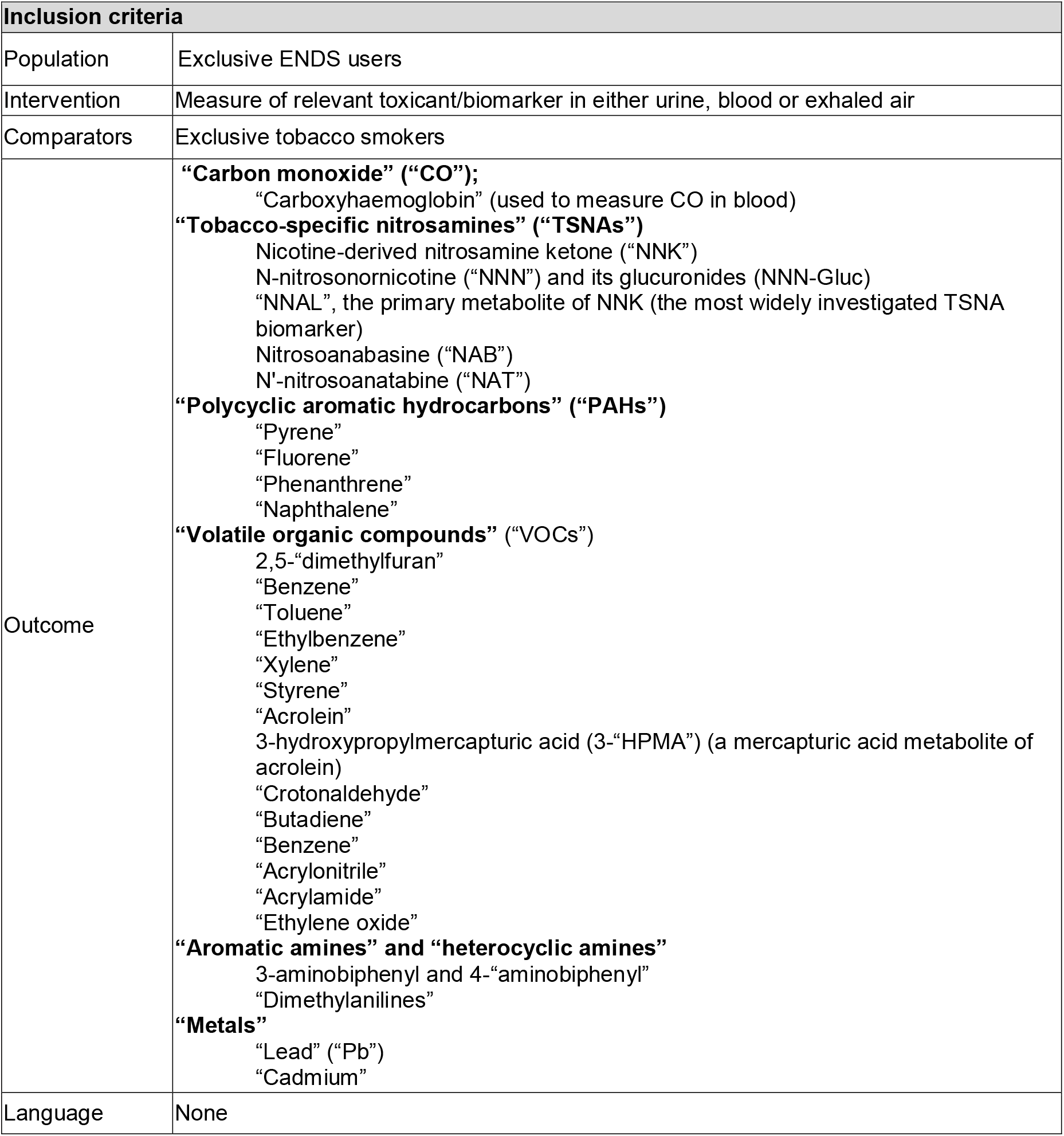
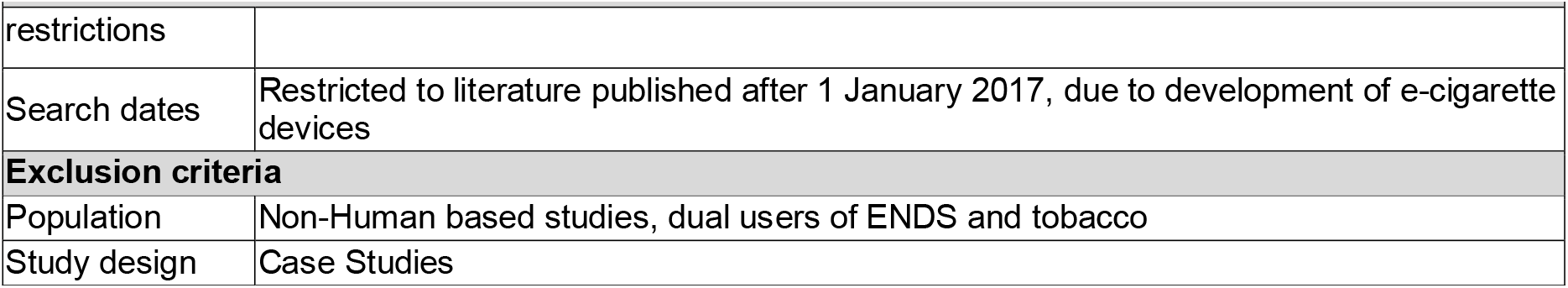
Literature search criteria (PICO framework)

**Supplementary Table 2:** Database Search Terms for PubMed. (Search date 1 September 2020)

| ID | Search | Hits |
| --- | --- | --- |
| #1 | Carbon monoxide OR "CO" OR Carboxyhaemoglobin | 889,062 |
| #2 | "Tobacco-specific nitrosamin*" OR TSNA | 876 |
| #3 | "Nicotine-derived nitrosamine ketone" OR NNK | 1,413 |
| #4 | N-nitrosanornicotine OR NNN OR glucuronid* OR NNN-N-Gluc* | 48,961 |
| #5 | NNAL OR methylnitrosamino | 1,476 |
| #6 | Nitrosoanabasine OR NAB | 5,579 |
| #7 | N'-nitrosoanatabine OR N-nitrosoanatabine OR NAT | 207,991 |
| #8 | "Polycyclic aromatic hydrocarbon*" OR PAH OR Pyrene OR Fluorene OR Phenanthrene OR Naphthalene | 306,959 |
| #9 | "Volatile organic compound*" OR VOC | 18,799 |
| #10 | "2,5-dimethylfuran" OR dimethylfuran OR DMF | 17,546 |
| #11 | Benzene OR Toluene OR Ethylbenzene OR Xylene OR Styrene OR Acrolein | 104,255 |
| #12 | "3-hydroxypropylmercapturic acid" OR 3-HPMA OR HPMA | 1,028 |
| #13 | Crotonaldehyde OR Butadiene OR Benzene OR Acrylonitrile OR Acrylamide OR "Ethylene oxid*" | 82,155 |
| #14 | "Aromatic amine*" OR "heterocyclic amine*" | 5,995 |
| #15 | 3-aminobiphenyl OR 3-Aminobiphenyl OR Aminobiphenyl OR Dimethylaniline* | 2,867 |
| #16 | Metals or "metal and lead" OR "Pb" OR Cadmium OR "Cd" | 1,573,488 |
| #17 | #1 OR #2 OR #3 OR #4 OR #5 OR #6 OR #7 OR #8 OR #9 OR #10 OR #11 OR #12 OR #13 OR #14 OR #15 OR #16 | 3,014,484 |
| #18 | ENDS OR vape* OR vapi* OR vapo* OR e-cig* | 132,178 |
| #19 | tobacco OR nicotine OR smok* | 407,192 |
| #20 | "2017/01/01"[Date - Publication] : "2020/08/25"[Date - Publication] | 4,590,129 |
| #21 | #17 AND #18 AND #19 AND #20 | 475 |

**Supplementary Table 3:** Google Scholar searches. (search date 1 September 2020. Restricted to articles published between 2017 and 2020 and sorted by relevance. The first ten pages of each search was assessed due to the large number of hits).

| ID | Search | Hits |
| --- | --- | --- |
| #1 | (Carbon OR monoxide OR Carboxyhemoglobin OR Carboxyhaemoglobin) AND ((tobacco OR nicotine OR smoke OR smoking) AND (ENDS OR vape OR vapor OR vaping OR vaporised OR vaporised OR e-cig OR e-cigarette)) | 23,200 |
| #2 | (Tobacco-specific OR nitrosamine OR TSNA) AND ((tobacco OR nicotine OR smoke OR smoking) AND (ENDS OR vape OR vapor OR vaping OR vaporised OR vaporised OR e-cig OR e-cigarette)) | 3,600 |
| #3 | (Nicotine-derived nitrosamine ketone OR NNK) AND ((tobacco OR nicotine OR smoke OR smoking) AND (ENDS OR vape OR vapor OR vaping OR vaporised OR vaporised OR e-cig OR e-cigarette)) | 257 |
| #4 | (N-nitrosornicotine OR NNN OR glucuronide OR NNN-N-Gluc) AND ((tobacco OR nicotine OR smoke OR smoking) AND (ENDS OR vape OR vapor OR vaping OR vaporised OR vaporised OR e-cig OR e-cigarette)) | 9,830 |
| #5 | (NNAL OR methylnitrosamino) AND ((tobacco OR nicotine OR smoke OR smoking) AND (ENDS OR vape OR vapor OR vaping OR vaporised OR vaporised OR e-cig OR e-cigarette)) | 20,800 |
| #6 | (Nitrosoanabasine OR NAB) AND ((tobacco OR nicotine OR smoke OR smoking) AND (ENDS OR vape OR vapor OR vaping OR vaporised OR vaporised OR e-cig OR e-cigarette)) | 3,370 |
| #7 | (N'-nitrosoanatabine OR N-nitrosoanatabine OR NAT) AND ((tobacco OR nicotine OR smoke OR smoking) AND (ENDS OR vape OR vapor OR vaping OR vaporised OR vaporised OR e-cig OR e-cigarette)) | 21,200 |
| #8 | ("Polycyclic aromatic hydrocarbon" OR PAH OR Pyrene OR Fluorene OR Phenanthrene OR Naphthalene) AND ((tobacco OR nicotine OR smoke OR smoking) AND (ENDS OR vape OR vapor OR vaping OR vaporised OR vaporised OR e-cig OR e-cigarette)) | 15,700 |
| #9 | ("Volatile organic compound*" OR VOC) AND ((tobacco OR nicotine OR smoke OR smoking) AND (ENDS OR vape OR vapor OR vaping OR vaporised OR vaporised OR e-cig OR e-cigarette)) | 9,420 |
| #10 | (2,5-dimethylfuran OR dimethylfuran OR DMF) AND ((tobacco OR nicotine OR smoke OR smoking) AND (ENDS OR vape OR vapor OR vaping OR vaporised OR vaporised OR e-cig OR e-cigarette)) | 4,120 |
| #11 | (Benzene OR Toluene OR Ethylbenzene OR Xylene OR Styrene OR Acrolein) AND ((tobacco OR nicotine OR smoke OR smoking) AND (ENDS OR vape OR vapor OR vaping OR vaporised OR vaporised OR e-cig OR e-cigarette)) | 17,000 |
| #12 | (3-hydroxypropylmercapturic OR 3-HPMA OR HPMA) AND ((tobacco OR nicotine OR smoke OR smoking) AND (ENDS OR vape OR vapor OR vaping OR vaporised OR vaporised OR e-cig OR e-cigarette)) | 314 |

**Supplementary Figure 1:**
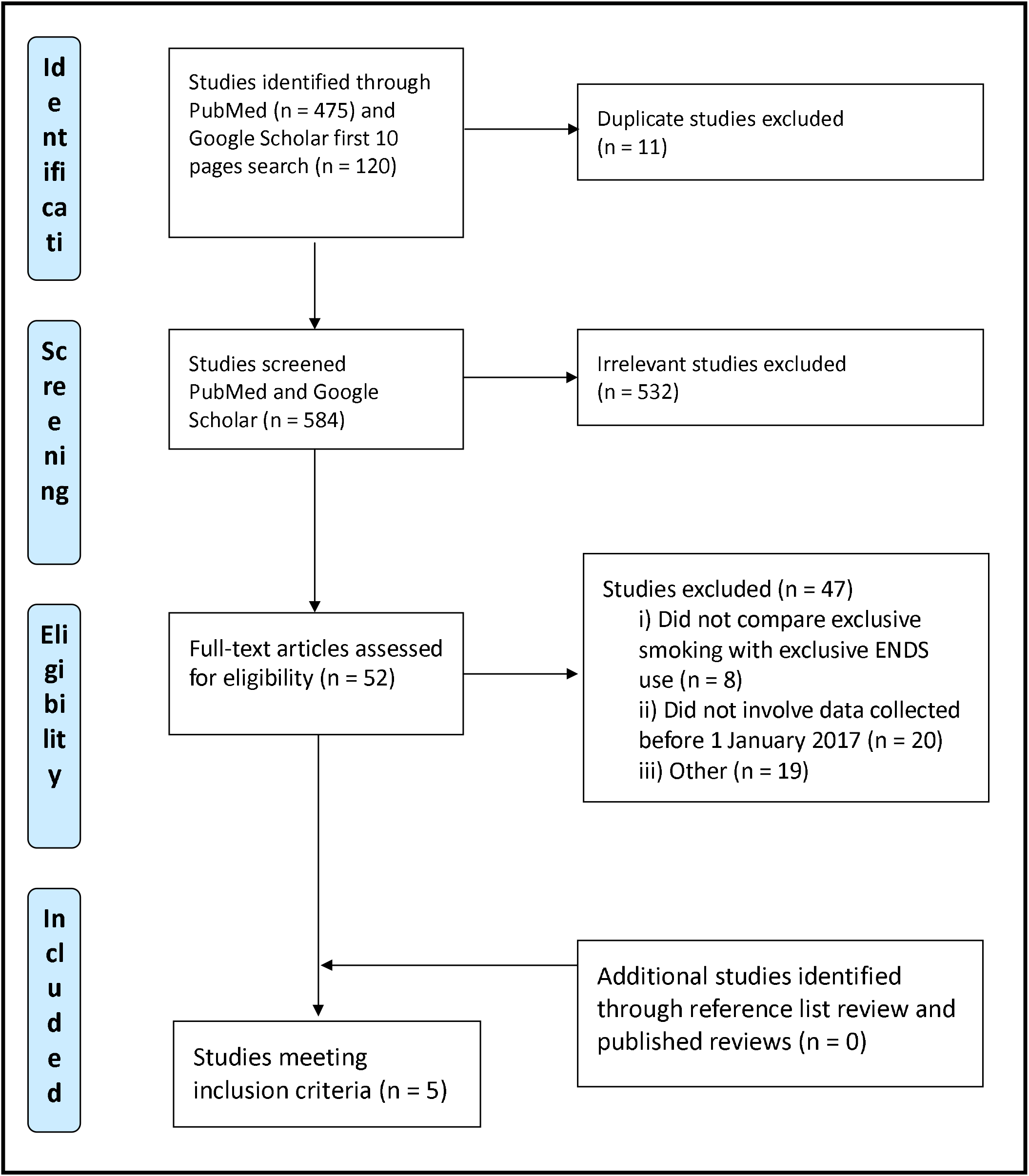
PRISMA diagram for the literature search.

**Supplementary Table A4:** Additional details on the five identified studies used in the analysis.

| Study | Dates of data collection | ENDS products used by participants | Additional details and comments |
| --- | --- | --- | --- |
| Boykan et al 2019 [2] | April 2017 to April 2018 | Mainly “pods” (78% of past-week daily users) | This was a cross-sectional study comparing exclusive tobacco users and exclusive e-cigarette users (albeit with no data on previous smoking status in the latter group). It involved those aged 12 to 21 years old and was a convenience sample of 3 outpatient clinics (Stony Brook, New York State, USA). The very much lower urinary cotinine levels in exclusive e-cigarette users vs exclusive smokers suggest low levels of unreported dual use in the e-cigarette users. However, these low cotinine levels also suggest low intensity of ENDS use that might be atypical relative to typical (and more intensive) ENDS use. Hence the biomarker levels for ENDS users in this study may be an underestimate of the true risk. A particular strength of this study was that it collected both self-reported marijuana use (ever, past 30 days, past 7 days) and the associated urinary biomarker data (tetrahydrocannabinolic acid: THCA). As a result, our analyses could use the results from those smokers and ENDS users who were not also marijuana users. |
| Hatsukami 2020 [3] | Study was initiated in 2014 but with the last follow-up on 2 December, 2018 | “Vuse Solo” | This study was a randomised controlled trial (RCT) in a US adult daily smoker population. Participants were randomized to different products, including one arm to complete substitution of cigarettes with ENDS. “Participants were incentivized for protocol compliance”. The biomarker results were based on the 8 week point. |
| Jay et al 2020 [4] | Between February and July 2018 | The JUUL nicotine-salt pod system (4 flavours: Virginia Tobacco; mint; mango; and crème) | This was a “randomized, open-label, parallel-cohort, confinement study of healthy adult smokers”. Randomisation was to 4 different flavours of e-cigarettes, to continuation of usual brand, or to cigarette abstinence. The population was based in Lincoln, New England, USA. This study was “sponsored by JUUL Labs, Inc.” (the manufacturer of the ENDS used in the study). Due to the short study time (5 days) we did not include the results for NNAL (owing to the long half-life of this biomarker – see main text). |
| Nga et al 2020 [5] | Not stated but the year on the grant number and on the ethics application was “2019” | Aspire AVP AIO Kit 700 mAh battery having a 1.2 X coil with e-liquid “Liquideo Evolution” | This was a “quasi experimental study” with convenience sampling (i.e., of staff, supporting staff or patients visiting the Oral Health Center, International Medical University Kuala Lumpur, Malaysia). Participants (all smokers at 10+ cigarettes per day for 5 years) were allowed to select the products being studied (ENDS or a heated tobacco product). Most participants were male (87%) and of Chinese ethnicity (51%). Participants were instructed to remain abstinent from cigarette smoking 12 hours before the experiment (and this was verified with eCO measurement). |
| Oliveri et al 2020 [6] | Between January 2017 and June 2017 | “Own brand of EVP” (e-vapour product). Results indicated 70 used tank-based | This was a cross-sectional, observational study. Participants were 30–65 years of age. Recruitment focused on the “four regions defined by the US Census Bureau to provide a geographically diverse study population.” The study was conducted by employees of Altria Client Services LLC (Altria is a tobacco company). |
|  |  | products and 62 used cartridge-based products | "Participants were instructed to use their own brand of EVP or conventional cigarettes ad libitum throughout study duration (approximately 30 days from electronic consent and online questionnaire completion to biological specimen collection)." The exclusive ENDS users "identified themselves as former smokers (minimum of 10 cigarettes per day for at least 10 years) and were exclusively using EVPs for at least 6 months and no other tobacco- or nicotine-containing products during that period." Exclusive adult smokers were "individuals who currently smoked 10 or more conventional cigarettes and have smoked 10 or more cigarettes per day for at least 10 years and did not use other tobacco or nicotine-containing products (including EVPs) in the past 30 days." |

**Supplementary Table A5:** Identified studies of acrolein (3-HPMA) in non-smokers and smokers of relevance to estimating the typical intake derived from non-smoking sources (ordered by decreasing number of study participants as bracketed)

| Study | Level in non-smokers [A] | Level in smokers [B] | % of [A] relative to [B] | Details |
| --- | --- | --- | --- | --- |
| Alwis et al 2015 [7] | 219<br>(n=2296) | 1089<br>(n=570) | 20.1% | Median levels in µg/g creatinine. From NHANES survey data for the USA. Given the size of this study it was the relative value from this that we used for the calculations in Table 1. |
| Scherer et al 2007 [8] | 337<br>(n=100) | 1297<br>(n=194) | 26.0% | Mean levels in µg/24h. Involved separate groups. The study was financially supported by a subsidiary of the German cigarette manufacturers' association. |
| Frigerio et al 2020 [9] | 160.6<br>(n=38) | 1301.2<br>(n=22) | 12.3% | Median levels in µg/g creatinine. Involved separate groups. This was a study of an occupational group in an industry with exposure to volatile chemicals. |
| Carmella et al 2009 [10] | 1500<br>(n=17) | 10,020<br>(n=17) | 15.0% | Mean levels in nmol/24h at day 56 after quitting smoking (i.e., the same group of individuals was studied). |
| D'Ruiz et al 2016 [11] | 228.8<br>(n=13) | 2004.1<br>(n=13) | 11.4% | Mean levels in µg/24h. Involved the same group – at start of study and after 5 days of abstinence. This study was funded by a fully owned subsidiary of Imperial Brands plc, and the manufacturer of the ENDS used in this study. |
| Lorkiewicz et al 2019 [12] | 294.3<br>(n=12) | 544.7<br>(n=8) | 54.0% | Mean levels in (ng/mg creatinine). Involved separate groups. Of note was that this study used first generation "cig-a-likes". |

